# Mapping the antigenic diversification of SARS-CoV-2

**DOI:** 10.1101/2022.01.03.21268582

**Authors:** Karlijn van der Straten, Denise Guerra, Marit J. van Gils, Ilja Bontjer, Tom G. Caniels, Hugo D.G. van Willigen, Elke Wynberg, Meliawati Poniman, Judith A. Burger, Joey H. Bouhuijs, Jacqueline van Rijswijk, Wouter Olijhoek, Marinus H. Liesdek, A. H. Ayesha Lavell, Brent Appelman, Jonne J. Sikkens, Marije K. Bomers, Alvin X. Han, Brooke E. Nichols, Maria Prins, Harry Vennema, Chantal Reusken, Menno D. de Jong, Godelieve J. de Bree, Colin A. Russell, Dirk Eggink, Rogier W. Sanders

## Abstract

Large-scale vaccination campaigns have prevented countless hospitalizations and deaths due to COVID-19. However, the emergence of SARS-CoV-2 variants that escape from immunity challenges the effectiveness of current vaccines. Given this continuing evolution, an important question is when and how to update SARS-CoV-2 vaccines to antigenically match circulating variants, similar to seasonal influenza viruses where antigenic drift necessitates periodic vaccine updates. Here, we studied SARS-CoV-2 antigenic drift by assessing neutralizing activity against variants-of-concern (VOCs) of a unique set of sera from patients infected with a range of VOCs. Infections with D614G or Alpha strains induced the broadest immunity, while individuals infected with other VOCs had more strain-specific responses. Omicron BA.1 and BA.2 were substantially resistant to neutralization by sera elicited by all other variants. Antigenic cartography revealed that Omicron BA.1 and BA.2 are antigenically most distinct from D614G, associated with immune escape and likely requiring vaccine updates to ensure vaccine effectiveness.

## Main text

The COVID-19 pandemic, caused by the SARS-CoV-2 virus, represents an enormous threat to human health and a burden to healthcare systems and economies worldwide. The unprecedented rapid development of efficacious vaccines fuelled hope of curtailing this pandemic and permitting a return to a society without societal restrictions. However, genetic drift of SARS-CoV-2 resulted in the emergence of multiple variants of concern (VOCs) with a higher transmissibility compared to the ancestral strain, and that challenge the effectiveness of public health measures, vaccines and/or therapeutics (World Health Organization, 2021). Based on this definition, the WHO designated the Alpha (Pango lineage B.1.1.7), Beta (B.1.351), Gamma (P.1), Delta (B.1.617.2) and Omicron (B.1.1.529, sublineages BA.1 and BA.2) variants as VOCs. The Alpha, Beta, Gamma and Delta VOCs have approximately 7 to 12 mutations in the Spike protein (S), while Omicron BA.1 with 34 mutations, of which 3 deletions, and BA.2 with 28 mutations, differ substantially more from the ancestral strain (Figure 1A)(World Health Organization, 2021). Approximately half of Omicron’s S mutations are located in the Receptor Binding Domain (RBD) and eight mutations in the N-terminal domain (NTD), the two most important antigenic site of S. Indeed, sera from COVID-19 patients infected with the ancestral strain and sera from vaccinees show up to 7-fold and 4-fold reductions in neutralization activity against Beta and Gamma, while 20 to 40-fold reductions are observed against Omicron BA.1 (Caniels et al., 2021; Garcia-Beltran et al., 2021; van Gils et al., 2022; Wilhelm et al., 2021).

**Figure 1.**
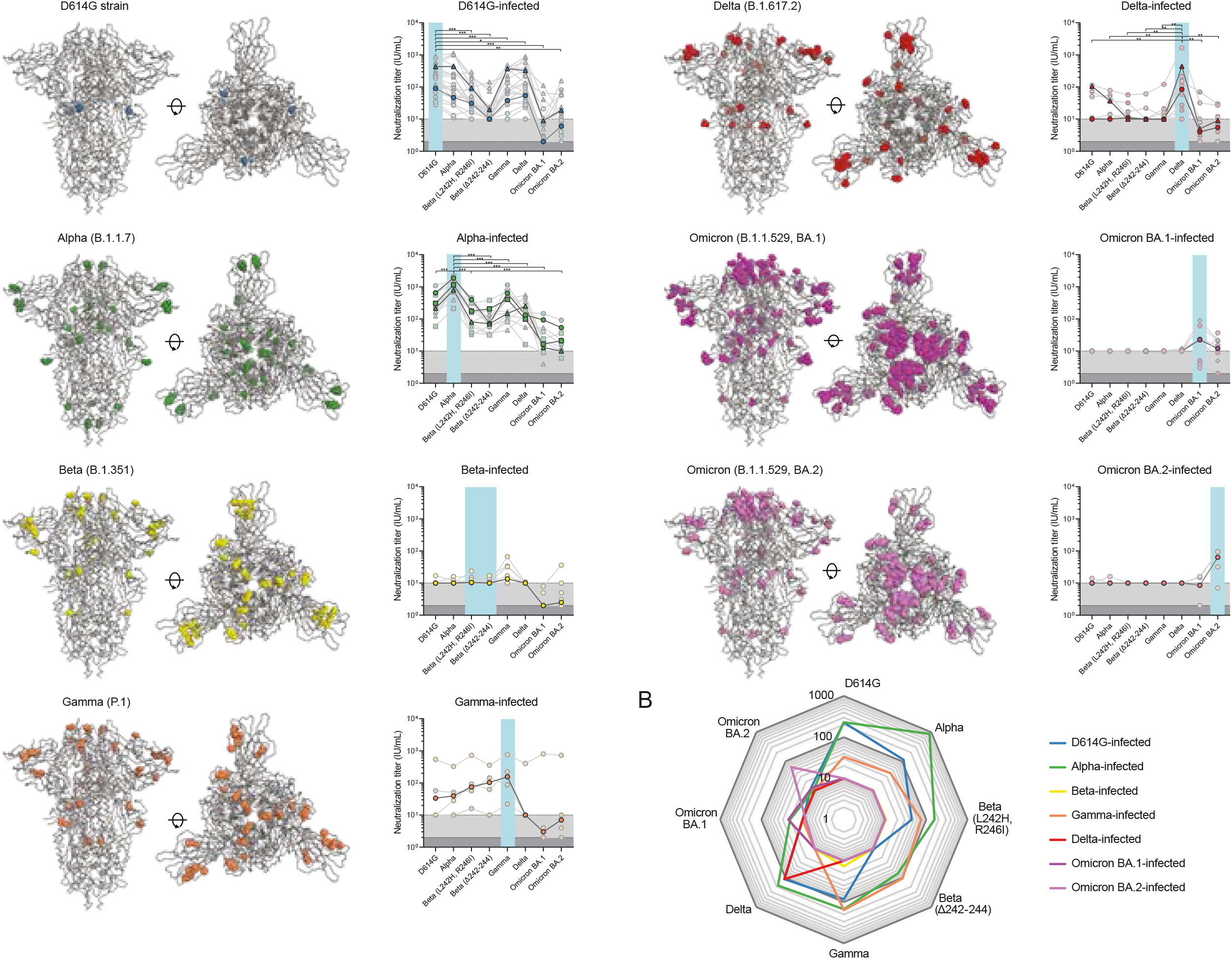
SARS-CoV-2 genetic diversity and neutralization. **A**. Molecular models of SARS-CoV-2 S, highlighting the locations of mutations in the D614G strain (blue), Alpha (green), Beta (yellow), Gamma (orange), Delta (red), Omicron BA.1 (magenta) and Omicron BA.2 (pink) variants. Midpoint neutralization titres against the VOCs in International Units per mL (IU/mL). The individuals are grouped per VOC and plotted accordingly. Median neutralization titres are highlighted while the individual points are depicted with higher transparency. The light grey bar (10 IU/ml) indicates the neutralization cut-off for all strains except Omicron (cut-off 2 IU/mL, dark grey bar). Non-hospitalized patients are indicated with dots and hospitalized patients with triangles. The individuals that were infected with an Alpha strain that also included the E484K mutation are indicated in green squares. The two individuals in the Omicron BA.1 group that may have been infected with BA.2 instead of BA.1 are indicated in magenta diamonds (see also Table S1). The homologous neutralization is highlighted using a light blue bar. The Wilcoxon signed rank test with Benjamini Hochberg correction was used to compare cross-neutralization titres with the homologous neutralization (see Table S3A for exact p-values). Only statistically significant differences are indicated. * = p< 0.005, ** = p<0.01, **** = p<0.0001. **B**. Spider plot of the median neutralization titre (IU/mL) of each group against all VOCs. A cut-off of 10 IU/mL is used for all strains.

However, the precise antigenic relationships between these VOCs are only starting to become clear. Understanding the differences between the serological antibody responses elicited by these variants is important to assess the risk of re-infections after natural infection and breakthrough infections after vaccination. For seasonal influenza viruses, this type of antigenic data is combined with virus genetic and epidemiological data to quantify the evolution of the virus and guide annual updates of the seasonal influenza virus vaccines. Antigenic cartography can be used to visualize antigenic relationships between viral variants (Fonville et al., 2014; Smith et al., 2004) and is routinely used in influenza virus vaccines strain selection. Until recently, antigenic cartography for SARS-CoV-2 has only been applied to cohorts of COVID-19 patients with uncertainty about their history of previous SARS-CoV-2 exposure and COVID-19 vaccinations, and without the usage of Omicron infected human data (Liu et al., 2021; Mykytyn et al., 2022; Wilks et al., 2022). Here, we studied the (cross-) neutralizing antibody responses in sera from a well-defined population of convalescent individuals with a sequence confirmed, or high likelihood of, primary infection by the D614G, Alpha, Beta, Gamma, Delta or Omicron BA.1 or BA.2 variants and used this data as input for antigenic cartography to map the antigenic evolution of SARS-CoV-2.

We collected and analysed a unique set of serum samples from 66 COVID-19 patients with a PCR-confirmed primary SARS-CoV-2 infection who did not receive any COVID-19 vaccinations. Blood was drawn 3 to 11 weeks after symptom onset (median 40 days, range 24 to 75 days), which corresponds with the peak of the antibody response (Table 1 and Table S1)(Long et al., 2020). In total, n=20 D614G, n=11 Alpha, n=8 Beta, n=4 Gamma, n=11 Delta, n=8 Omicron BA.1 and n=4 Omicron BA.2 infected participants were included. Of these participants, 39 had a sequence-confirmed VOC infection. The other 27 participants met our inclusion criteria of a high likelihood of VOC infection (see STAR Methods section and Table S1), of which 20 participants were assumed to be infected with the D614G strain as they were sampled before the emergence of any VOC in the Netherlands, but after D614G became dominant (Korber et al., 2020).

**Table 1.** Sociodemographics and clinical characteristics. A summary of the convalescent SARS-CoV-2 patients included in this study. Table S1 contains a more comprehensive overview per individual.

| Variant of concern | N= (%) | Age (years)<br>median (range) | Male | Hospital<br>admission | Sequence<br>confirmed | Time since<br>symptom onset<br>(days)<br>median (range) |
| --- | --- | --- | --- | --- | --- | --- |
| **D614G** | 20 (30%) | 53 (22-74) | 9 (45%) | 9 (45%) | 0 (0%) | 38 (30-45) |
| **Alpha** | 11 (17%) | 48 (25-76) | 6 (55%) | 9 (82%) | 11 (100%) | 39 (24-58) |
| Alpha + E484K | 2/11 | 62 (48-76) | 0 | 2/2 | 2/2 | 41 (24-58) |
| **Beta** | 8 (12%) | 41 (18-53) | 3 (38%) | 0 | 7 (88%) | 39 (30-63) |
| **Gamma** | 4 (6%) | 35 (27-10) | 2 (50%) | 0 | 4 (100%) | 40 (38-55) |
| **Delta** | 11 (17%) | 31 (19-63) | 6 (55%) | 1 (9%) | 10 (91%) | 46 (36-52) |
| **Omicron BA.1** | 8 (12%) | 31 (21-45) | 4 (50%) | 0 | 3 (38%) | 44 (25-75) |
| **Omicron BA.2** | 4 (6%) | 51 (31-55) | 4 (100%) | 0 | 4 (100%) | 40 (39-41) |
| Total | 66 | 41 (18-76) | 34 (52%) | 21 (32%) | 39 (59%) | 40 (24-75) |

We assessed the neutralizing capacity of the convalescent sera in a lentiviral-based pseudovirus neutralization assay against the D614G strain, the Alpha, two Beta, the Gamma, Delta and Omicron BA.1 and BA.2 variants (Figure 1A). The two Beta subvariants differ from each other in the NTD, where one Beta subvariant (L242H, R246I) is based on a very early available sequence while the other (Δ242-244) is retrospectively more representative for the predominant circulating strains.

The highest neutralization titres were generally measured against the homologous virus, as might be expected (Figure 1A and Figure S1A). Only the Beta infected participants showed higher cross-neutralization titres against the Gamma variant compared to homologous neutralization, which is in line with other research(Wilks *et al*., 2022). This might be explained by the shared RBD mutations, as the RBDs of these variants only differ by one amino acid (K417N in Beta *versus* K417T in Gamma). Our analyses suffer somewhat from a disbalance in hospitalized versus non-hospitalized patients between the different VOC groups(Table 1). However, when comparing only non-hospitalized patients which generally have lower antibody levels compared to hospitalized patients, patients infected with the Alpha variant showed the strongest homologous neutralization (1881 IU/mL, range 1658 to 2103 IU/mL), followed by individuals infected with the Gamma variant (median of 156 IU/mL, range 22 to 761 IU/mL), the D614G strain (median of 90 IU/mL, range 28 to 237 IU/mL) the Delta variant (median of 85 IU/mL, range 10 to 1635 IU/mL), the Omicron BA.2 (median of 64 IU/mL, range 10 to 95 IU/mL) and the Omicron BA.1 variant (median of 23 IU/mL, range 10 to 90 IU/mL) (Figure S1A). By contrast, none of the Beta infected participants showed substantial homologous neutralization against either Beta subvariants (Figure 1A, Figure S2).

Overall, the VOCs differed in their capacity to induce cross-neutralizing antibodies. Individuals infected with the Alpha variant induced the broadest response, followed by D614G strain-infected, Gamma-infected and Delta-infected patients (Figure 1A, Figure 1B and Figure S1B), though there was substantial heterogeneity within all groups. Notably, none of the patients infected with the Beta, Omicron BA.1 or Omicron BA.2 variants showed substantial cross-neutralization activity.

Reductions in neutralizing activity against the two Omicron variants were substantial in all groups (Figure 1A and 1B). Omicron neutralization dropped below the limit of detection (10 IU/mL or an ID_50_ of 100) in 44/66 of the studied individuals for BA.1 and 37/66 for BA.2. The median fold-reduction of Omicron BA.1 neutralization versus homologous neutralization was 9-fold (range 1 to 93-fold) when considering all patients, 10-fold (range 3 to 93-fold) for patients infected with a D614G strain, 52-fold (range 11 to 89-fold) for Alpha, 6-fold (range 1 to 22-fold) for Gamma, and 6-fold (range 1 to 51-fold) for Delta infected patients. The median fold-reduction of Omicron BA.2 neutralization versus homologous neutralization was 5.4-fold (range -3.7 to 134-fold) when considering all patients, 8-fold (range 3 to 47-fold) for patients infected with D614G strain, 68-fold (range 18 to 134-fold) for Alpha, 6-fold (range 1 to 22-fold) for Gamma, and 6-fold (range 1 to 60-fold) for Delta infected patients.

To explore the antigenic relationships between the VOCs, we used the neutralization data to construct a SARS-CoV-2 antigenic map (Figure 2A). In this map, homologous sera tend to cluster around the infecting strain, reflecting that homologous neutralization is dominant. The D614G and Alpha viruses cluster tightly together in the centre of the map, while the Beta (L242H, R246I), Gamma, and Delta variants all lie within 2 antigenic units (1 unit = 2-fold change in neutralization titre) of the D614G strain suggesting a high degree of antigenic similarity. For influenza viruses, variants are considered to be antigenically similar in case of antigenic distances below 3 antigenic units, i.e. an 8-fold change in neutralization titre, and different when above this threshold(Barr et al., 2014; Prevention, 2021). By analogy, the D614G, Alpha, Beta (L242H, R246I), Gamma and Delta variants belong to one antigenic cluster. Interestingly, the Beta (Δ242-244) subvariant is antigenically more distinct from the D614G strain compared to Beta (L242H, R246I) (e.g. 3 to 4 units), implying that the deletion at region 242-244 has a substantial effect on antigenicity and illustrates the importance of the NTD as target of neutralizing antibodies and/or in modulating antigenicity of other domains by allosteric means. The distance between the main antigenic cluster and Omicron BA.1 and BA.2 variants is more than 4 antigenic units (>16-fold change in neutralization) implying that Omicron BA.1 and BA.2 are the antigenically most distinct SARS-CoV-2 variants (Figure 2A). One caveat is that it is unclear whether 2-fold changes in pseudovirus neutralization titres are directly comparable to 2-fold changes in hemagglutination inhibition assay titres used to define different antigenic clusters of influenza viruses. However, the change in neutralization between Omicron BA.1 and BA.2 and other variants of SARS-CoV-2, including the D614G strain, is striking.

**Figure 2.**
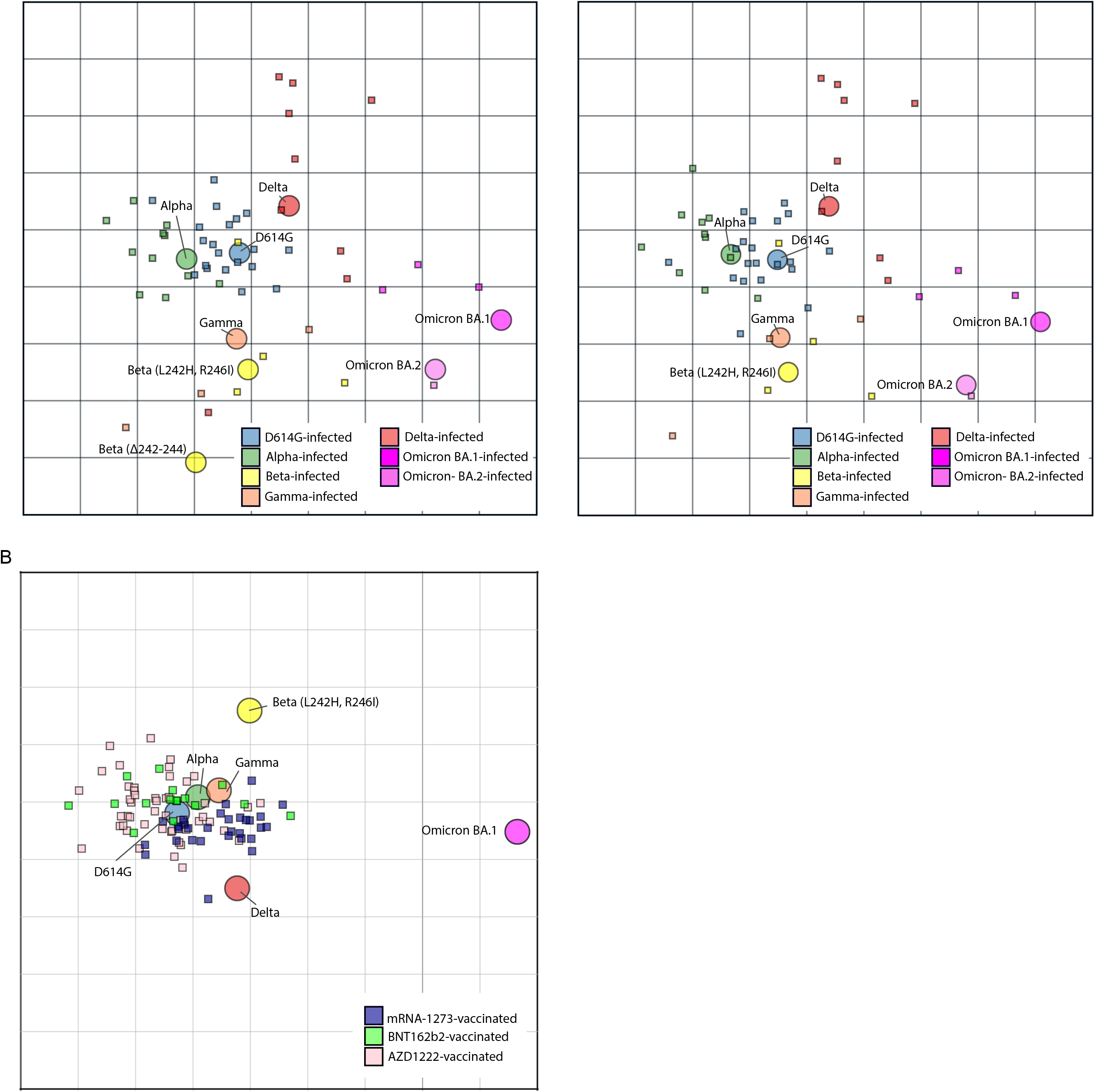
SARS-CoV-2 antigenic cartography. **A**. Antigenic map of SARS-CoV-2 VOCs based on convalescent SARS-CoV-2 infection sera. SARS-CoV-2 variants are shown as circles and sera are indicated as squares. Each square corresponds to sera of one individual and is coloured by the infecting SARS-CoV-2 variant. Both axes of the map are antigenic distance and each grid square (1 antigenic unit) represents a two-fold change in neutralization titre. The distance between points in the map can be interpreted as a measure of antigenic similarity, where the points more closely together show higher cross-neutralization and are therefore antigenically more similar. The left panel included both Beta subvariants used in this study. The right panel is without the Beta (Δ242-244) subvariant **B**. Antigenic map of SARS-CoV-2 VOCs based on post-vaccination sera from individuals without prior SARS-CoV-2 infections. Each serum is coloured by the vaccine that individual received.

We next used neutralization data from sera of 109 COVID-19 naïve vaccinees receiving either two Moderna (mRNA-1273, n=30), Pfizer/BioNTech (BNT162b2, n=49), or AstraZeneca (AZD1222, n=30) vaccines, which are all based on the ancestral S sequence to generate a second antigenic map (van Gils *et al*., 2022). This map (Figure 2B) agreed well with the infectee maps (Figure 2A), and corroborated that Omicron BA.1 represents a distinct antigenic variant from viruses currently included in vaccines. Interestingly, while the distributions of sera from recipients of different vaccines overlap, there is a skew of sera of mRNA-1273 vaccinees towards Omicron BA.1, suggesting small differences in antigen stimulation among vaccine formulations considered here.

We have started to define the antigenic SARS-CoV-2 landscape after two years of antigenic drift, which should inform risk assessment of re-infections as well as strain selection for COVID-19 vaccine updates. We can draw several conclusions. First, homologous neutralization was usually stronger than heterologous neutralization. Second, heterologous responses were broadest and most potent in individuals infected with Alpha and D614G strains, while infection with Delta resulted in narrow-specificity responses. In addition, the individuals infected with the Beta and Omicron BA.1 variant, and to lesser extent Omicron BA.2 infected individuals, developed weak neutralizing responses against any VOC, including the homologous strains, suggesting that the S proteins of Beta and both Omicron variants are less immunogenic compared to the S of other VOCs. Interestingly, the weak homologous and cross-neutralization levels of Beta variant infected individuals are in contrast with the higher titres found by others(Cele et al., 2022; Liu *et al*., 2021; Rossler et al., 2022; Wilks *et al*., 2022). It is unlikely that this weak homologous neutralization is caused by a sequence mismatch between the strains causing the infection and the sequence used in our pseudoviruses, as both Beta subvariant pseudoviruses escape homologous neutralization of Beta sera, and the reduction of cross-neutralization of sera elicited by other VOCs against the Beta variants are in line with previous studies (Cele *et al*., 2022; Liu *et al*., 2021; Rossler *et al*., 2022; Wilks *et al*., 2022). One contributing factor for these low (cross-)neutralization titres by the Beta variant includes cohort specific differences between studies, which is, however, hard to verify due to limited patient characteristic and demographic information available in publications. The weak homologous and cross-neutralization of Omicron BA.1 variant infected individuals is in line with other pre-print data(Mykytyn *et al*., 2022). Third, the D614G and Alpha strains are at the centre of our antigenic map, which supports the use of the current COVID-19 vaccines based on the ancestral strain, in case of circulation of the Alpha, Beta (L242H, R246I), Gamma and Delta variants. Our data suggest that updated vaccines based on the Beta (L242H, R246I) or Delta variants would not have been appreciably more effective than the ancestral virus-based vaccines. However, the substantial reduction of neutralization in all groups against the Beta Δ242-244, but especially against the Omicron variants indicates a high risk of re-infections and vaccine breakthrough cases when exposed to these VOCs. The long antigenic distance between Omicron variants and the preceding variants in the antigenic map indicates that the current high rates of Omicron infections are at least partially associated with immune escape and that a vaccine update is required. While finishing this study, several other efforts to antigenically characterize VOCs became available (Mykytyn *et al*., 2022; Wilks *et al*., 2022). Our antigenic cartography is largely in accordance with these other studies.

As in the case of seasonal influenza viruses, the prospect of SARS-CoV-2 becoming an endemic virus with recurring outbreaks implies the need for surveillance of antigenic drift and possibly yearly administration of updated vaccines, especially for individuals at risk for severe COVID-19. Antigenic cartography efforts such as those presented here, can inform future vaccine updates.

## Supporting information

Supplemental Table 1. Sociodemographics and clinical characteristics

## Data Availability

All data produced in the present study are available upon reasonable request to the authors.

## Acknowledgements

We thank all public health services (GGD) in the Netherlands for their help in contacting participants. We are also thankful to the study personnel and the participants of the COSCA, RECoVERED and the S3-study for their contribution to this research.

## Author Contributions

Conceptualization, K.vdS., D.G., M.J.vG., C.R., C.A.R., D.E., R.W.S.,

Methodology, K.vdS., D.G., M.J.vG., C.A.R., D.E., R.W.S.,

Validation, K.vdS., D.G., M.J.vG., I.B.,

Formal Analysis, K.vdS., A.X.H., B.E.N., C.A.R.,

Investigation, I.B., M.Po., J.A.B., J.H.B., J.vR., W.O.,

Resources, K.vdS., I.B., H.D.G.vW., E.W., M.H.L., A.H.A.L., B.A., J.L.S., M.K.B., M.Pr., H.V., C.R., M.D.dJ, G.J.dB,

Writing – original draft, K.vdS., D.G., A.X.H., C.A.R., D.E., R.W.S.,

Writing – review & editing, M.J.vG, I.B., T.G.C., H.D.G.vW., E.W., M.Po., J.A.B., J.H.B., J.vR., W.O., M.H.L., A.H.A.L., B.A., J.J.S., M.K.B., B.E.N., M.Pr., H.V., C.R., M.D.dJ., G.J.dB.

Visualization, K.vdS.,T.G.C., A.X.H., C.A.R., R.W.S.,

Supervision, C.A.R., D.E., R.W.S.,

Project administration, K.vdS., D.G.,

Funding acquisition, J.J.S., M.K.B., A.X.H., M.P., M.D.dJ., G.J.dB., C.A.R., R.W.S.,

## STAR METHODS

### KEY RESOURCES TABLE

| REAGENT or RESOURCE | SOURCE | IDENTIFIER |
| --- | --- | --- |
| DMEM media | Gibco | Cat#: 11966025 |
| Opti-MEM I reduced serum media | Gibco | Cat#: 15392402 |
| Phosphate-buffered saline (PBS) | Gibco | Cat#: 15326239 |
| **Bacterial and virus strains** |  |  |
| Chemically competent DH5α *Escherichia coli* | Thermo Fisher Scientific | Cat#: 12879416 |
| **Biological samples** |  |  |
| Human sera, convalescent SARS-CoV-2 | This study | N/A |
| Human sera, post-COVID-19 vaccination | (van Gils *et al.*, 2022) | N/A |
| **Chemicals, peptides, and recombinant proteins** |  |  |
| FastDigest SacI | Thermo Fisher Scientific | Cat#: 10324720 |
| FastDigest ApaI | Thermo Fisher Scientific | Cat#: 10450280 |
| Polyethylenimine hydrochloride (PEI) MAX | PolySciences | Cat#: 24765-1 |
| Trypsin-EDTA | Gibco | Cat#: 25-200-056 |
| GlutaMAX | Gibco | Cat#: 35050061 |
| Glycylglycine, 99+% | Acros Organics/Thermo Scientific | Cat#: 120141000 |
| Magnesium sulfate heptahydrate, 99.5%, for analysis, Thermo Scientific™ | Acros Organics/Thermo Scientific | Cat#: AC21311500 |
| EGTA (ethylene glycol bis(2-aminoethyl ether)-N,N,N',N'-tetraacetic acid) ≥97%, Ultra Pure Grade | VWR | Cat#: 0732-100G |
| Triton™ X-100 (Electrophoresis), Fisher BioReagents™ | Fisher BioReagents | Cat#: BP151-500 |
| Poly-L-Lysine Hydrobromide | Sigma-Aldrich | Cat#: P1399 |
| **Critical commercial assays** |  |  |
| Gibson Assembly | New England BioLabs | E5510S |
| QuikChange Site-Directed Mutagenesis Kit | Agilent Technologies | Cat#: 200523 |
| **Experimental models: Cell lines** |  |  |
| HEK293T cells | American Type Culture Collection | CRL-11268 |
| HEK-293T-hACE2 | (Schmidt *et al.*, 2020) | RRID:CVCL_A7UK |
| **Oligonucleotides** |  |  |
| SARS-CoV-2 D614G spike gene fragment | Integrated DNA Technologies | GenBank:MT449663.1 Mutations in this paper |
| SARS-CoV-2 Alpha spike gene fragment | Integrated DNA Technologies | GenBank:MT449663.1 Mutations in this paper |
| SARS-CoV-2 Beta spike gene fragment | Integrated DNA Technologies | GenBank:MT449663.1 Mutations in this paper |
| SARS-CoV-2 Gamma spike gene fragment | Integrated DNA Technologies | GenBank:MT449663.1<br>Mutations in this paper |
| SARS-CoV-2 Delta spike gene fragment | Integrated DNA Technologies | GenBank:MT449663.1<br>Mutations in this paper |
| SARS-CoV-2 Omicron BA.1 spike gene fragment | Integrated DNA Technologies | GenBank:MT449663.1<br>Mutations in this paper |
| SARS-CoV-2 Omicron BA.2 spike gene fragments | Integrated DNA Technologies | GenBank:MT449663.1<br>Mutations in this paper |
| **Recombinant DNA** |  |  |
| pCR3 SARS-CoV-2-S<sub>Δ19</sub> expression plasmid | GenBank | ID: MT449663.1 |
| pHIV-1<sub>NL43</sub> ΔEnv-NanoLuc reporter virus plasmid | (Schmidt *et al.*, 2020) |  |
| **Software and algorithms** |  |  |
| ACMACS antigenic cartography software | https://acmacs-web.antigenic-cartography.org | N/A |
| GraphPad Prism 8.3.0 | GraphPad | RRID:SCR_002798;<br>http://www.graphpad.com/ |
| Microsoft Excel | Microsoft | RRID:SCR_016137;<br>https://www.microsoft.com/en-gb/ |
| **Other** |  |  |
| Nano-Glo Luciferase Assay System | Promega | Cat#: N1130 |
| GloMax system | Turner BioSystems | Cat#: 9101-002 |

## RESOURCE AVAILABILITY

### Lead contact

Further information and requests for resources and reagents should be directed to and will be fulfilled by the lead contact, Rogier W. Sanders

### Materials availability

This study did not generate new unique reagents.

### Data and code availability

Neutralisation and patient characteristic data have been deposited as supplementary tables within this manuscript (Table S1 and Table S2) and are publicly available as of the date of publication. Other accession numbers are listed in the key resources table. This paper does not report original code. Any additional information required to reanalyse the data reported in this paper is available from the lead contact upon request.

## EXPERIMETNAL MODEL AND SUBJECT DETAILS

### Study population

66 adults (aged 18 to 76) with a PCR proven primary SARS-CoV-2 infection were included in the COSCA-study (NL 73281.018.20) or the RECOVERED study (NL73759.018.20) between June 2020 and April 2022 at Amsterdam UMC and via the Dutch national SARS-CoV-2 sequence surveillance program as described previously(Grobben et al., 2021; Wynberg et al., 2021). In short, 3-11 weeks after symptom onset, blood, patient demographics, time between symptom onset and sampling, and admission status were collected (Table 1 and Table S1). The diagnostic oropharyngeal swab was available for 39 participants and were used to determine the SARS-CoV-2 strain causing the infection. The remaining 27 SARS-CoV-2 infected participants fell within the following inclusion criteria: (1) ≥95% of circulating strains at time of symptom onset belonged the suspected VOC of infection or (2) ≥75% of circulating strains at the time of symptom onset belonged the suspected VOC of infection AND a household member had a concurrent sequence confirmed infection with that particular VOC. Prevalence data of CoVariants.org and the National Institute for Public Health and the Environment were used to determine the current prevalence of a VOC (CoVariants, 2022; Rijksinstituut voor Volksgezondheid en Milieu (RIVM)). Most individuals of which no sequence confirmation of the infected strain was available, were presumed to be infected with de D614G variant (n=20) as they were sampled before the emergence of any VOC in the Netherlands and after D614G became predominant in the Netherlands(Korber *et al*., 2020). More details about the remaining n=7 individuals can be found in the Table S1. The two Omicron individuals that may have been infected by either BA.1 or BA.2 are indicated as diamonds in all graphs. Two of the individuals infected with an Alpha strain harbouring the E484K mutation are indicated as squares in all graphs.

Neutralization data on COVID-19 naive vaccinee sera were kindly provided by the S3-study of the Amsterdam UMC, The Netherlands(NL73478.029.20) (van Gils *et al*., 2022). In short, post-vaccination sera was obtained approximately four weeks after the second doses of either Moderna (mRNA-1273), Pfizer/BioNTech (BNT162b2), or AstraZeneca (AZD1222). Post-vaccination serum after Janssen (Ad26.COV2.S) were excluded from analysis because they did not have enough non-threshold titres to be included in the map.

All above mentioned studies were conducted at the Amsterdam University Medical Centres, the Netherlands, and approved by the local ethical committees. All individuals provided written informed consent before participating.

### Pseudovirus design

The D614G strain and the Alpha pseudovirus constructs contained the following mutations: D614G in D614G strain; deletion (Δ) of H69, V70 and Y144, N501Y, A570D, D614G, P681H, T716I, S982A, and D1118H in Alpha. The two Beta subvariants differ from each other in the NTD region 242-246, where one Beta subvariant (L242H, R246I) is based on a very early available sequence while the other (Δ242-244) is retrospectively more representative for the predominant circulating strains. These two Beta pseudovirus constructs contain therefore the following mutations: L18F, D80A, D215G, L242H, R246I, K417N, E484K, N501Y, D614G, and A701V in Beta (L242H, R246I); L18F, D80A, D215G, Δ242-244, K417N, E484K, N501Y, D614G, and A701V in Beta (Δ242-244). Only the D614G infected individuals showed statistically significant reduced neutralization against the Beta (Δ242-244) subvariant compared to the Beta (L242H, R246I) subvariant (Figure S2). The Gamma pseudovirus constructs contained the following mutations: L18F, T20N, P26S, D138Y, R190S, K417T, E484K, N501Y, D614G, H655Y, and T1027I in Gamma; This Gamma pseudovirus construct differs from the predominant strain in that it lacks a V1176F back bone mutation. However, it is not likely that this mutation, positioned at the S2 domain of the S, will affect escape of neutralization substantially. The Delta and Omicron BA.1 and BA.2 pseudovirus constructs contained the following mutations: T19R, G142D, E156G, Δ157-158, L452R, T478K, D614G, P681R and D950N in Delta; A67V, Δ69-70, T95I, G142D, Δ143-145, Δ211, L212I, ins214EPE, G339D, S371L, S373P, S375F, K417N, N440K, G446S, S477N, T478K, E484A, Q493K, G496S, Q498R, N501Y, Y505H, T547K, D614G, H655Y, N679K, P681H, N764K, D796Y, N856K, Q954H, N969K, L981F in Omicron BA.1; and T19I, L24S, Δ125/127, G142D, V213G, G339D, S371F, S373P, S375F, T376A, D405N, R408S, K417N, N440K, S477N, T478K, E484A, Q493R, Q498R, N501Y, Y505H, D614G, H655Y, N679K, P681H, N764K, D796Y, Q954H, N969K in Omicron BA.2. The Omicron BA.1 strain used here harbors a Q493K mutation, while the predominant Omicron BA.1 harbors a Q493R mutation. This mutation did not impacted neutralization of several monoclonal SARS-CoV-2 antibodies tested (data not shown). The spike constructs were ordered as gBlock gene fragments (Integrated DNA Technologies) and cloned SacI and ApaI in the pCR3 SARS-CoV-2–S_Δ19_ expression plasmid (GenBank: MT449663.1) using Gibson Assembly (Thermo Fisher Scientific). The pseudovirus constructs were made using the QuikChange Site-Directed Mutagenesis Kit (Agilent Technologies) and verified by using Sanger sequencing. Pseudoviruses were procedures by cotransfecting HEK293T cells (American Type Culture Collection, CRL-11268) with the pCR3 SARS-CoV-2-S_Δ19_ expression plasmid and the pHIV-1_NL43_ ΔEnv-NanoLuc reporter virus plasmid. Transfection takes place in cell culture medium (DMEM), supplemented with 10% fetal bovine serum, penicillin (100 U/ml), streptomycin (100 g/ml). Medium is refreshed once 6-8 hours after transfection. 48 hours after the transfection, cell supernatants containing the pseudovirus were harvested and stored at -80 °C until further use.

## METHOD DETAILS

### SARS-CoV-2 pseudovirus neutralization assay

The pseudovirus neutralization assay was performed as described previously(Caniels *et al*., 2021). Shortly, HEK293T/ACE2 cells were kindly provided by P. Bieniasz(Schmidt *et al*., 2020) were seeded at a density of 20,000 cells per well in a 96-well plate coated with poly-lysine (50 ug/ml) 1 day before the start of the neutralization assay. The next day, heat-inactivated sera samples were in triplicate serial diluted in threefold steps, starting at 1:20 dilution to test for Omicron BA.1 and BA.2 pseudovirus neutralization and 1:100 for all the other variants. Sera was diluted in cell culture medium (DMEM), supplemented with 10% fetal bovine serum, penicillin (100 U/ml), streptomycin (100 g/ml), and GlutaMAX (Gibco), mixed in a 1:1 ratio with pseudovirus, and incubated for 1 hour at 37°C. These mixtures were then added to the cells in a 1:1 ratio and incubated for 48 hours at 37°C, and lysis buffer was added. The luciferase activity in cell lysates was measured using the Nano-Glo Luciferase Assay System (Promega) and GloMax system (Turner BioSystems). Relative luminescence units were normalized to those from cells infected with SARS-CoV-2 pseudovirus in the absence of sera. The inhibitory neutralization titres (ID_50_) were determined as the serum dilution at which infectivity was inhibited by 50%, using a nonlinear regression curve fit (GraphPad Prism software version 8.3). The International Standard for anti-SARS-CoV-2 immunoglobulins provided by the WHO(Kristiansen et al., 2021) were used to convert the ID_50_ values into International Units per milliliters (IU/mL). Samples with IU/mL titres <10 were defined as having undetectable neutralization against the D614G, Alpha, Beta, Gamma and Delta variant. For Omicron BA.1 and BA.2 neutralization, the start-dilution of 1:20 enables a cut-off of <2 IU/mL for all samples except for some Alpha infected individuals. A limited amount of sera was available from the Alpha infected individuals, resulting in a start dilution of 1:100 of n=7 samples against all variants including Omicron BA.1 and BA.2. Neutralization data points of two Alpha infected individuals against BA.1 and BA.2 were excluded from Figure 1A because a neutralization titres <10IU/mL (Table S2). This exclusion did not impact the statistics as written below because a general cut-off of <10IU/mL were used for neutralization against any variant, including Omicron BA.1 and BA.2.

### Antigenic cartography

Antigenic maps were constructed as previously described (Fonville *et al*., 2014; Smith *et al*., 2004) using the antigenic cartography software from https://acmacs-web.antigenic-cartography.org. In brief, this approach to antigenic mapping uses multidimensional scaling to position antigens (viruses) and sera in a map to represent their antigenic relationships. The maps here relied on the SARS-CoV-2 post-infection serology data and post-vaccination serology data shown in Figure 1A and Table S2. The positions of antigens and sera were optimized in the map to minimise the error between the target distances set by the observed pairwise virus-serum combinations in the pseudovirus assay described above and the resulting computationally derived map. Maps were constructed in 2, 3, 4, and 5 dimensions to investigate the dimensionality of the antigenic relationships. Both the convalescent (Figure 2A) and post-vaccination datasets (Figure 2B) were strongly two-dimensional with only small improvements in residual mean squared error of the maps as map dimensionality increased.

## QUANTIFICATION AND STATISTICAL ANALYSIS

Data visualization and statistical analyses were performed in GraphPad Prism software (version 8.3). Spider plots (Figure 1B) were made in Excel 2016. The antigenic maps were produced using the antigenic cartography software mentioned above. Wilcoxon signed rank test with Benjamini Hochberg correction was used to compare cross-neutralization titres with the homologous neutralization (Figure 1A). Mann-Whitney test was used for non-paired group comparisons (Figure S1). All statistics mentioned here were performed by using a general neutralization cut-off of 10IU/mL against any variant of SARS-CoV-2.

