## Supplemental Table 1. Sociodemographics and clinical characteristics for "Mapping the antigenic diversification of SARS-CoV-2"

**This PDF file includes:**

Supplementary Figure 1 (Figure S1)
Supplementary Figure 2 (Figure S2)
Supplementary Table 1 and corresponding legend (Table S1)
Supplementary Table 3 and corresponding legend (Table S3)

**Seperate excel file:**

Supplementary Table 2 and corresponding legend

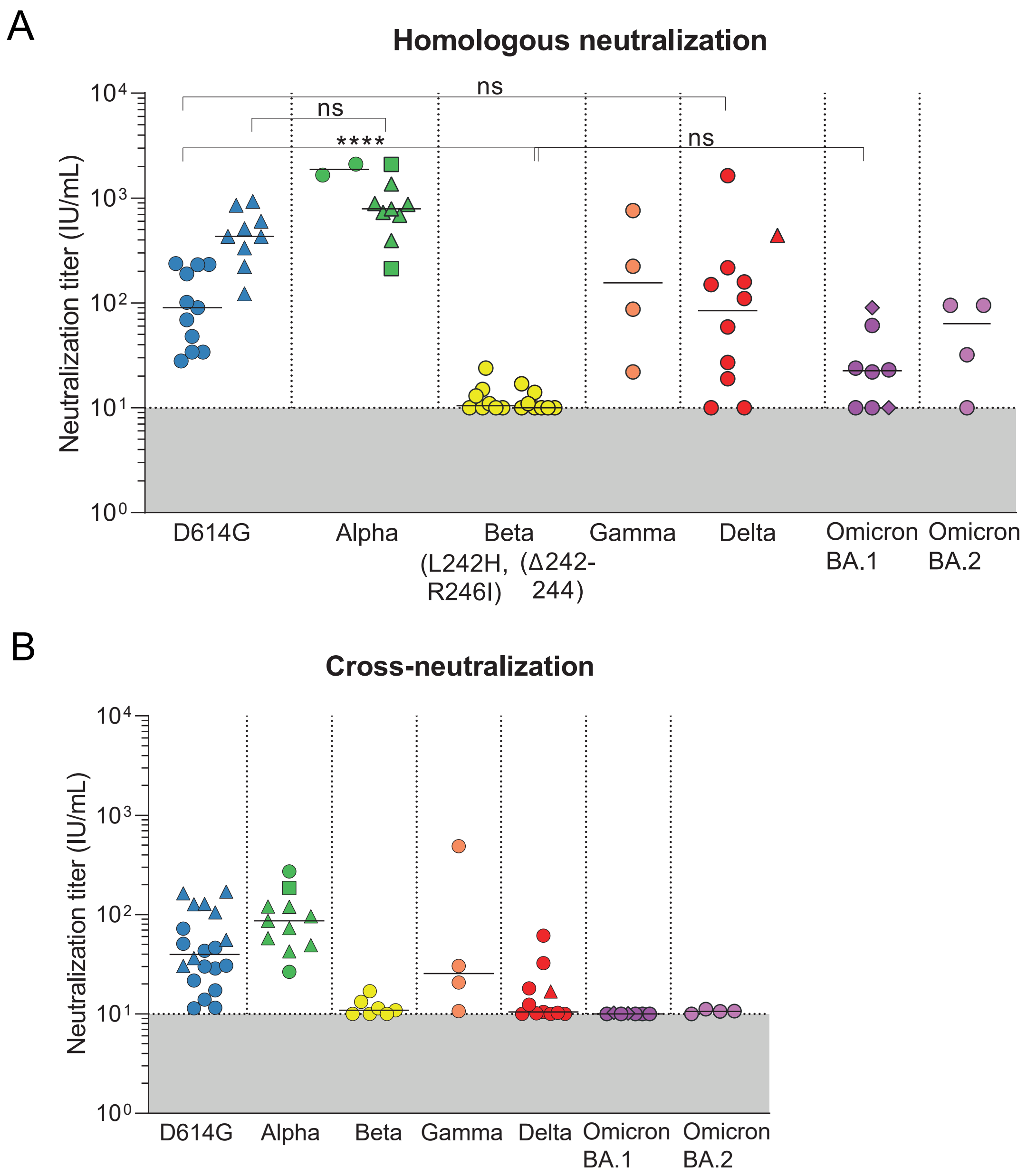


**Supplementary Figure 1. Homologous and cross-neutralization after SARS-CoV-2 infection. A.** Midpoint neutralization titres against the VOCs in International Units per mL (IU/mL). The individuals are grouped per VOC they were infected with and plotted accordingly. Non-hospitalized patients are indicated with dots and hospitalized patients with triangles. The individuals that were infected with an Alpha variant that also included the E484K mutation are indicated in green squares. The two individuals in the Omicron BA.1 group that may have been infected with BA.2 instead of BA.1 are indicated in magenta diamonds (see also Table S1). A MannWhitney test is used to test for differences between group medians (black lines). ns= non-significant, **** = p<0.0001. (see Table S3B for exact p-values)**B.** Cross-neutralization is expressed as the geometric mean of the neutralization titres against all VOCs except the autologous strain in IU/mL. A cut-off of 10IU/mL is used for all neutralization titres, as indicated by the grey bar.

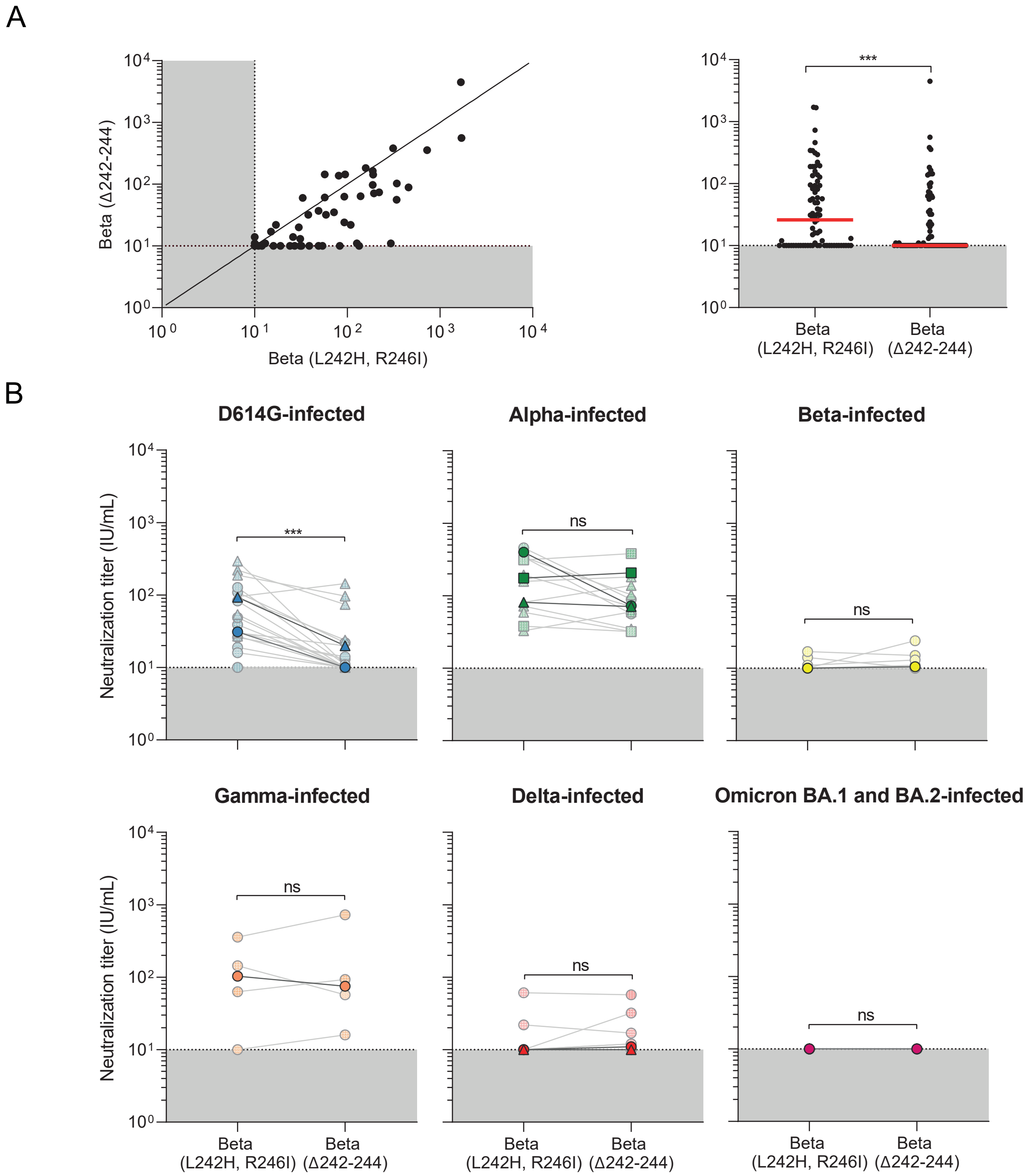


**Supplementary Figure 2. Neutralization against Beta subvariant pseudoviruses. A.** The left panel shows the correlation between the midpoint neutralization titres against both Beta subvariants used in this research, expressed in International Units per mL (IU/mL). In the right panel we studied the difference in neutralization titre against both Beta subvariants using a Wilcoxon signed rank test. Median neutralization titres are depicted as red bars. *** = p<0.001. **B.** Midpoint neutralization titres against both Beta subvariants in International Units per mL (IU/mL). The individuals are grouped per VOC they were infected with and plotted accordingly. Non-hospitalized patients are indicated with dots and hospitalized patients with triangles. The individuals that were infected with an Alpha variant that also included the E484K mutation are indicated in green squares. Median neutralization titres were compared using a Wilcoxon signed rank test. ns= non-significant, *** = p< 0.001.

**Supplemental Table 1. Sociodemographics and clinical characteristics**. Overview of sociodemographics and clinical characteristics of all convalescent SARS-CoV-2 patients included in this study.

| Pat ID | Variant of Concern (VOC) | Likelyhood of VOC infection | | Age at sampling | Gender | WHO severity score | Fever 1=yes 2=no | Admission 1= yes 2= no | IC 1= yes 2= no | Duration ofadmission (days) | Days between symptom onset and sampling |
|---|---|---|---|---|---|---|---|---|---|---|---|
| | | Sequence confirmed | Highly likely based on: | | | | | | | | |
| **COSCA-020** | D614G | | D614G was the only circulating variant | 46-55 | Female | 2 | 1 | 2 | - | - | 36 |
| **COSCA-021** | D614G | | D614G was the only circulating variant | 18-25 | Female | 2 | 2 | 2 | - | - | 31 |
| **COSCA-022** | D614G | | D614G was the only circulating variant | 18-25 | Male | 1 | 1 | 2 | - | - | 42 |
| **COSCA-023** | D614G | | D614G was the only circulating variant | 46-55 | Female | 2 | 1 | 2 | - | - | 43 |
| **COSCA-024** | D614G | | D614G was the only circulating variant | 18-25 | Female | 1 | 1 | 2 | - | - | 30 |
| **COSCA-025** | D614G | | D614G was the only circulating variant | 18-25 | Male | 1 | 2 | 2 | - | - | 30 |
| **COSCA-026** | D614G | | D614G was the only circulating variant | 66-75 | Male | 1 | 1 | 2 | - | - | 35 |
| **COSCA-028** | D614G | | D614G was the only circulating variant | 36-45 | Female | 1 | 1 | 2 | - | - | 30 |
| **COSCA-033** | D614G | | D614G was the only circulating variant | 46-55 | Female | 1 | 2 | 2 | - | - | 33 |
| **COSCA-034** | D614G | | D614G was the only circulating variant | 26-35 | Male | 1 | 1 | 2 | - | - | 40 |
| **COSCA-112** | D614G | | D614G was the only circulating variant | 46-55 | Male | 3 | 1 | 1 | 2 | 4 | 39 |
| **COSCA-113** | D614G | | D614G was the only circulating variant | 66-75 | Male | 4 | 1 | 1 | 1 | 9 | 43 |
| **COSCA-114** | D614G | | D614G was the only circulating variant | 56-65 | Female | 2 | 1 | 2 | - | - | 35 |
| **COSCA-115** | D614G | | D614G was the only circulating variant | 56-65 | Male | 3 | 1 | 1 | 2 | 6 | 40 |
| **COSCA-116** | D614G | | D614G was the only circulating variant | 56-65 | Female | 3 | 1 | 1 | 2 | 3 | 31 |
| **COSCA-117** | D614G | | D614G was the only circulating variant | 46-55 | Female | 3 | 1 | 1 | 2 | 8 | 38 |
| **COSCA-118** | D614G | | D614G was the only circulating variant | 56-65 | Male | 3 | 1 | 1 | 2 | 2 | 37 |
| **COSCA-119** | D614G | | D614G was the only circulating variant | 56-65 | Female | 2 | 1 | 1 | 2 | 1 | 40 |
| **COSCA-120** | D614G | | D614G was the only circulating variant | 66-75 | Male | 2 | 1 | 1 | 2 | 5 | 45 |
| **COSCA-123** | D614G | | D614G was the only circulating variant | 36-45 | Female | 2 | 1 | 1 | 2 | 4 | 39 |
| **COSCA-303** | B.1.1.7 | Sequence confirmed | | 66-75 | Male | 2 | 1 | 2 | - | - | 27 |
| **GGDVIS3343** | B.1.1.7 | Sequence confirmed | | 18-25 | Female | 1 | 1 | 2 | - | - | 44 |
| **VUVIS6992** | B.1.1.7 | Sequence confirmed | | 66-75 | Male | 2 | 1 | 1 | 2 | 4 | 32 |
| **AMCVIS1585** | B.1.1.7 | Sequence confirmed | | 26-35 | Female | 3 | 1 | 1 | 2 | 3 | 51 |
| **AMCVIS1871** | B.1.1.7 | Sequence confirmed | | 26-35 | Male | 2 | 1 | 1 | 2 | 4 | 40 |
| **AMCVIS2084** | B.1.1.7 | Sequence confirmed | | 26-35 | Male | 3 | 1 | 1 | 2 | 9 | 39 |
| **AMCVIS3484** | B.1.1.7 | Sequence confirmed | | 26-35 | Female | 2 | 1 | 1 | 2 | 6 | 32 |
| **AMCVIS5722** | B.1.1.7 | Sequence confirmed | | 46-55 | Male | 3 | 2 | 1 | 2 | 9 | 35 |
| **AMCVIS9584** | B.1.1.7 | Sequence confirmed | | 46-55 | Male | 2 | 1 | 1 | 2 | 3 | 48 |
| **COSCA-316** | B.1.1.7 + E484K | Sequence confirmed | | >75 | Female | 3 | 1 | 1 | 2 | 8 | 24 |
| **COSCA-320** | B.1.1.7 + E484K | Sequence confirmed | | 46-55 | Female | 4 | 2 | 1 | 1 | 11 | 58 |
| **COSCA-301** | B.1.351 | Sequence confirmed | | 46-55 | Female | 1 | 1 | 2 | - | - | 41 |
| **COSCA-331** | B.1.351 | Sequence confirmed | | 18-25 | Female | 1 | 2 | 2 | - | - | 55 |
| **COSCA-336** | B.1.351 | Sequence confirmed | | 36-45 | Male | 1 | 2 | 2 | - | - | 42 |
| **COSCA-337** | B.1.351 | Sequence confirmed | | 36-45 | Female | 1 | 1 | 2 | - | - | 63 |
| **COSCA-305** | B.1.351 | Sequence confirmed | | 46-55 | Female | 1 | 2 | 2 | - | - | 30 |
| **COSCA-306** | B.1.351 | Sequence confirmed | | 46-55 | Male | 1 | 2 | 2 | - | - | 30 |
| **COSCA-307** | B.1.351 | | 75% of circulating strains belonged to the Beta variant and a household member had a concurrent sequence confirmed infection | 18-25 | Female | 1 | 1 | 2 | - | - | 30 |
| **COSCA-308** | B.1.351 | Sequence confirmed | | 18-25 | Male | 1 | 1 | 2 | - | - | 36 |
| **COSCA-309** | P.1 | Sequence confirmed | | 26-35 | Female | 1 | 2 | 2 | - | - | 41 |
| **COSCA-310** | P.1 | Sequence confirmed | | 36-45 | Male | 1 | 2 | 2 | - | - | 38 |
| **COSCA-324** | P.1 | Sequence confirmed | | 26-35 | Female | 2 | 1 | 2 | - | - | 38 |
| **COSCA-334** | P.1 | Sequence confirmed | | 36-45 | Male | 1 | 1 | 2 | - | - | 55 |
| **COSCA-321** | B.1.617.2 | Sequence confirmed | | 56-65 | Male | 1 | 2 | 2 | - | - | 43 |
| **COSCA-322** | B.1.617.2 | | 75% of circulating strains belonged to the Delta variant and a household member had a concurrent sequence confirmed infection | 18-25 | Male | 1 | 2 | 2 | - | - | 47 |
| **COSCA-323** | B.1.617.2 | Sequence confirmed | | 46-55 | Female | 1 | 1 | 2 | - | - | 46 |
| **COSCA-325** | B.1.617.2 | Sequence confirmed | | 46-55 | Male | 3 | 1 | 1 | 2 | 3 | 52 |
| **COSCA-327** | B.1.617.2 | Sequence confirmed | | 26-35 | Female | 1 | 1 | 2 | - | - | 47 |
| **COSCA-328** | B.1.617.2 | Sequence confirmed | | 36-45 | Male | 2 | 1 | 2 | - | - | 46 |
| **COSCA-329** | B.1.617.2 | Sequence confirmed | | 26-35 | Female | 1 | 2 | 2 | - | - | 44 |
| **COSCA-330** | B.1.617.2 | Sequence confirmed | | 18-25 | Female | 1 | 2 | 2 | - | - | 49 |
| **COSCA-332** | B.1.617.2 | Sequence confirmed | | 46-55 | Female | 1 | 2 | 2 | - | - | 36 |
| **COSCA-333** | B.1.617.2 | Sequence confirmed | | 26-35 | Male | 2 | 1 | 2 | - | - | 43 |
| **COSCA-335** | B.1.617.2 | Sequence confirmed | | 26-35 | Male | 2 | 1 | 2 | - | - | 37 |
| **COSCA-348** | Omicron BA.1 | | >95% of circulating strains belonged to BA.1 | 36-45 | Female | 1 | 1 | 2 | - | - | 39 |
| **COSCA-349** | Omicron BA.1 | | >95% of circulating strains belonged to BA.1 | 26-35 | Male | 1 | 2 | 2 | - | - | 42 |
| **COSCA-350** | Omicron BA.1 | Sequence confirmed | | 18-25 | Female | 1 | 1 | 2 | - | - | 45 |
| **COSCA-351** | Omicron BA.1 | | 75% of circulating strains belonged to the BA.1 variant and a household member had a concurrent sequence confirmed infection | 26-35 | Male | 1 | 2 | 2 | - | - | 68 |
| **COSCA-352** | Omicron BA.1/BA.2 | | 70% of circulating strains belonged to BA.1 and 30% belonged to BA.2 | 26-35 | Female | 1 | 2 | 2 | - | - | 25 |
| **COSCA-353** | Omicron BA.1/BA.2 | | 70% of circulating strains belonged to BA.1 and 30% belonged to BA.2 | 26-35 | Male | 2 | 1 | 2 | - | - | 31 |
| **COSCA-354** | Omicron BA.1 | Sequence confirmed | | 26-35 | Female | 1 | 2 | 2 | - | - | 75 |
| **COSCA-356** | Omicron BA.2 | Sequence confirmed | | 26-35 | Male | 1 | 1 | 2 | - | - | 40 |
| **COSCA-357** | Omicron BA.2 | Sequence confirmed | | 46-55 | Male | 1 | 2 | 2 | - | - | 39 |
| **COSCA-358** | Omicron BA.2 | Sequence confirmed | | 46-55 | Male | 1 | 1 | 2 | - | - | 41 |
| **COSCA-360** | Omicron BA.1 | Sequence confirmed | | 36-45 | Male | 1 | 1 | 2 | - | - | 59 |
| **COSCA-361** | Omicron BA.2 | Sequence confirmed | | 46-55 | Male | 1 | 1 | 2 | - | - | 40 |

**Supplementary Table 3. P-values of statistical analysis**

**Supplementary Table 3A. Statistical analysis of Figure 1A.** The Wilcoxon signed rank test with Benjamini Hochberg correction was used to compare cross-neutralization titres with the homologous neutralization.

| | **Infected individuals** | | | | | | |
|---|---|---|---|---|---|---|---|
| | **Ancestral** | **Alpha** | **Beta** | **Gamma** | **Delta** | **Omicron BA.1** | **Omicron BA.2** |
| **Ancestral** | | 0,0010 | 0,6000 | 0,1750 | 0,0046 | 0,0729 | 0,25 |
| **Alpha** | 0,1208 | | 0,6000 | 0,1750 | 0,0046 | 0,0729 | 0,25 |
| **Beta (L242H, R246I)** | 0,0002 | 0,0010 | | 0,1750 | 0,0046 | 0,0729 | 0,25 |
| **Beta (Δ242-244)** | 0,0002 | 0,0010 | | 0,3750 | 0,0059 | 0,0729 | 0,25 |
| **Gamma** | 0,0002 | 0,0010 | 0,3750 | | 0,0046 | 0,0729 | 0,25 |
| **Delta** | 0,0183 | 0,0010 | 0,3750 | 0,1750 | | 0,0729 | 0,25 |
| **Omicron BA.1** | 0,0002 | 0,0010 | 0,3750 | 0,3750 | 0,0046 | | 0,25 |
| **Omicron BA.2** | 0,0014 | 0,0010 | 0,8750 | 0,1750 | 0,0046 | 0,2188 | |

| | |
|---|---|
| ns | P > 0.05 |
| * | P ≤ 0.05 |
| ** | P ≤ 0.01 |
| *** | P ≤ 0.001 |

**Supplementary Table 3B. Statistical analysis of Figure S1A.** A MannWhitney test is used to test for differences between group medians.

| **P-values using Mann-Whitney U test** | **P-value** | |
|---|---|---|
| **Hospitalized patients** | | |
| D614G vs Alpha infected | 0,11 | ns |
| **Non-hospitalized** | | |
| D614G vs Beta(Δ242-244) infected | <0,0001 | **** |
| D614G vs Delta infected | 0,46 | ns |
| Beta (Δ242-244) vs Omicron BA.1 infected | 0,088 | ns |

**Supplementary Table 3C. Statistical analysis of Figure S2A.** A Wilcoxon signed rank test was used to study the differences in neutralization titres against both Beta subvariants.

| **P-values using Wilcoxon rank test** | | | | | | | |
|---|---|---|---|---|---|---|---|
| | Ancestral | Alpha | Beta | Gamma | Delta | Omicron BA.1 | Omicron BA.2 |
| Beta (L242H, R246I) vs Beta (Δ242-244) | 0,0004 | 0,21 | 0,875 | 0,625 | >0,99 | >0,99 | >0,99 |
